# Comparative evaluation of machine learning models versus TIMI score in ST-segment-elevation myocardial infarction patients

**DOI:** 10.1101/2024.05.16.24307511

**Authors:** Mohit D Gupta, Dixit Goyal, Shekhar Kunal, Manu Kumar Shetty, Girish MP, Vishal Batra, Ankit Bansal, Prashant Mishra, Mansavi Shukla, Aarti Gupta, Vanshika Kohli, Akul Chadha, Arisha Fatima, Subrat Muduli, Anubha Gupta, Jamal Yusuf

**Author notes:** Address for correspondence: Dr. Mohit Dayal Gupta Professor of Cardiology, Room no. 125, First floor, Academic block Department of Cardiology, Govind Ballabh Pant Institute of Postgraduate Medical Education & Research, New Delhi-110002, India, Ext-5125.

## Abstract

**Background:** Risk stratification is an integral component of ST-segment-elevation myocardial infarction (STEMI) management practices. This study aimed to derive a machine learning (ML) model for risk stratification and identification of factors associated with in-hospital and 30-day mortality in patients with STEMI and compare it with traditional TIMI score.

**Methods:** This was a single center prospective study wherein subjects >18 years with STEMI (n=1700) were enrolled. Patients were divided into two groups: training (n=1360) and validation dataset (n=340). Six ML algorithms (Extra Tree, Random Forest, Multiple Perceptron, CatBoost, Logistic Regression and XGBoost) were used to train and tune the ML model and to determine the predictors of worse outcomes using feature selection. Additionally, the performance of ML models both for in-hospital and 30-day outcomes was compared to that of TIMI score.

**Results:** Of the 1700 patients, 168 (9.88%) had in-hospital mortality while 30-day mortality was reported in 210 (12.35%) subjects. In terms of in-hospital mortality, Random Forest ML model (sensitivity: 80%; specificity: 74%; AUC: 80.83%) outperformed the TIMI score (sensitivity: 70%; specificity: 64%; AUC:70.7%). Similarly, Random Forest ML model (sensitivity: 81.63%; specificity: 78.35%; AUC: 78.29%) had better performance as compared to TIMI score (sensitivity: 63.26%; specificity: 63.91%; AUC: 63.59%) for 30-day mortality. Key predictors for worse outcomes at 30-days included mitral regurgitation on presentation, smoking, cardiogenic shock, diabetes, ventricular septal rupture, Killip class, age, female gender, low blood pressure and low ejection fraction.

**Conclusions:** ML model outperformed the traditional regression based TIMI score as a risk stratification tool in patients with STEMI.

## Introduction

ST segment elevation myocardial infarction (STEMI) is still the most dreaded manifestation of coronary artery disease having a high mortality despite all the significant advances such as reperfusion and percutaneous coronary interventions (PCI). Multiple registries have reported that in-hospital mortality in STEMI ranges between 7-10%^1^. Prompt diagnosis and management with proper risk stratification is crucial to reduce the mortality and morbidity associated with STEMI. There is a need to identify high-risk STEMI patients and triage them to various levels of hospital care along with judicious allocation of resources^2^. Over the past many years, multiple risk scores have been developed such as the Thrombolysis in Myocardial Infarction (TIMI) risk score^3^, Global Registry of Acute Coronary Events (GRACE) risk score^4^, Zwolle primary PCI risk score^5^, PAMI risk score^6^ and CADILLAC risk score^7^ to predict short and long-term mortality following STEMI with TIMI and GRACE being the most popular ones. These traditional risk scores were developed using Cox proportional regression models with pre-assumptions and researcher dependent feature selection comprising limited variables. This can lead to missing key features that might be contributing to mortality. Also, these scores have been developed more than two decades ago among Caucasian population and have not been systematically validated among South Asians which is a heterogenous population having inherent genetic variations in group of multi-ethnic communities^8^. In recent times, machine learning (ML) has emerged as a powerful tool to handle large clinical data while avoiding prior assumptions^9^. It has been used to create unbiased cardiovascular risk prediction models^10–12^. Interpretability methods including local interpretable model-agnostic explanations (LIME) for individual cases and the Shapley Additive explanations (ShAP) for both the population and individual cases can be combined with predictive accuracy of ML based models for better understanding of model’s decision-making process ^13,14^.

The present study aimed to develop and validate a customized ML based algorithm to predict in-hospital and 30-days mortality in patients presenting with STEMI in a large heterogeneous Indian population and compare it with the most widely used TIMI risk score. The designed ML model was validated on unseen dataset and the ML model’s decision was analyzed with population level ShAP plots for better understanding of the key features significantly impacting mortality.

## METHODS

### Study design and Study population

This was a single center, prospective, observational study comprising patients with STEMI who were admitted in a tertiary care hospital in New Delhi, India. All the patients aged 18 years and older presenting with a primary diagnosis of STEMI within 72 hours of symptom onset and consenting to be a part of the study were enrolled over a duration of 18 months w.e.f. 1^st^ August 2021 to 31^st^ January 2023. A diagnosis of STEMI was based on the European Society of Cardiology/American College of Cardiology Foundation/American Heart Association/World Heart Federation Task Force for the Fourth Universal Definition of Myocardial Infarction^15^. Patients with non-ST-segment myocardial infarction (NSTEMI), congenital heart disease, structural heart disease, pregnant females, and co-existing condition with life expectancy of less than six months were excluded. A total of 1700 patients fulfilling the inclusion criteria were enrolled for the study (Figure 1: central illustration). Written informed consent was taken from all the patients and proper approval for the study was taken from the institutional ethics committee (F.1/IEC/MAMC/85/04/2021/No.486).

**Figure 1:** Central illustration showing the overview of the methods used for data selection, use of machine learning (ML) algorithms, validation and the performance of ML model as well as TIMI score. (AUC: Area Under the Curve; ML: machine learning; MI: myocardial infarction; STEMI:ST-Segment Elevation Myocardial Infarction; TIMI: thrombolysis in myocardial infarction; mo: months; HF: heart failure; MR: mitral regurgitation; CAD: coronary artery disease; LVEF: left ventricular ejection fraction).

### Data collection and feature selection

The whole study data was divided into two groups: training dataset (80%) to develop the ML models and unseen data (20%) set for testing the ML model. The collected data ventricular septal rupture (VSR). In all the enrolled patients both in-hospital and 30-days all-cause mortality was calculated. Univariate and multivariate analysis were applied on the data set and the features that were significantly affecting the mortality were included to develop the ML model. These features included age, gender, diabetes mellitus, hypertension, smoking, body mass index (BMI), treatment delay > 1 hour, MI type on ECG (anterior wall, inferior wall, lateral wall, and posterior wall), heart rate > 100 beats per minute, systolic blood pressure < 90 mmHg, Killip class, cardiac status at presentation (cardiogenic shock and heart failure), time to treatment > 4 hours, LVEF (%) categories (20-25, 25-30, 30-35, 35-40, 40-45, and greater than 50), mitral regurgitation and VSR.

### Development and validation of ML models

The collected data underwent a specific preprocessing and refinement process to make it suitable for further analysis. Python’s Numpy and Pandas packages were used for data cleaning. The dataset was then split into two subsets: a training cohort, encompassing 80% of the patients and an unseen cohort comprising the remaining 20%. In order to ensure an equitable representation of patient outcomes (mortality), the data split was carefully designed to maintain an identical proportion of mortality cases in both the training and validation sets. Jupyter notebook in Python served as the platform for training the ML models and conducting testing. The dataset exhibited an expected inherent imbalance with a notably larger number of patients who survived compared to those who experienced mortality. Out of the 1,700 patients with STEMI, 168 were associated with in-hospital mortality and 210 were linked to 30-day mortality. In order to rectify this imbalance, resampling techniques were applied to the training set while preserving the validation set in its original form. The entire dataset was then evaluated using a total of 27 candidate features for in-hospital mortality and 30 features for 30-day mortality employing six distinct ML algorithms: Extra Tree, Random Forest, Multiple Perceptron, CatBoost, Logistic Regression and XGBoost.

### Performance measures and feature ranking

The performance of ML prediction models was evaluated by analyzing various metrics from the confusion matrix. These metrics included sensitivity, specificity, F1-score, receiver operating characteristic (ROC) area under the curve (AUC) and accuracy of the model. Sensitivity (true positive rate or recall) is the probability of identifying the true positives (model correctly identifying the patients with high risk of mortality). Specificity (true negative rate) on the other hand relates to the model’s ability to correctly identify the patients who will survive. Precision (positive predictive value) is the proportion of true positives divided by sum of true positives and false positives. F1 Score is the harmonic mean of precision and recall and is a better metric of incorrectly classified classes than Accuracy. The primary goal of the ML model was to provide highly precise predictions for in-hospital and 30-days mortality patients, with a strong emphasis on achieving a high true positive rate on the validation dataset. This is important in order to provide optimum medical care to the high-risk patients who require it promptly. SHAP interpretability plots were employed for the best-performing ML model. These plots are a part of Explainable AI which provides valuable insights into the reasoning behind the model’s predictions and outcomes.

### Comparison with TIMI score

TIMI score is a composite of 8 parameters with the score ranging from 0-14. TIMI score was calculated in all the patients according to original TIMI score developed by Morrow and colleagues^3^. The sensitivity, specificity, AUC and accuracy of the TIMI score, using a threshold score of 4 to distinguish between the mortality and survival classes was computed. Scores below 4 were categorized as the survival class, while scores equal to or greater than 4 were categorized as the mortality class. The performance metrics of the TIMI score were compared with those of the ML models.

### Statistical analysis

Continuous data were expressed as mean ± standard deviation (SD) and categorical data were denoted as proportions. A comparison of means of continuous variables was done using Student’s t-test or Mann-Whitney U test as appropriate, while Fisher exact test or χ2 test was used for categorical variables. A two-sided p-value of <0.05 was statistically significant. SPSS version 24.0 (IBM Corp, Armonk, NY) was used for statistical analysis.

## RESULTS

### Baseline characteristics

The mean age of the study population was 54.5±11.9 years. Most of them were males (1370 [80.6%]) and females represented (330[19.4%]) of the population. A total of 373 (22%) cases were diabetics and 534 (31%) were hypertensive. Out of all, 700 (41%) patients were smokers and approximately 67% of the population in our study was physically inactive (defined as less than 150 mins of aerobic exercise per week or less than 75 mins of vigorous exercise per week). Around 11% of patients presented with cardiogenic shock and only 11% of patients reached hospital within 1 hour of symptoms. Most common type of MI was anterior wall MI (56%) followed by IWMI (41%). A total of 1200 (70.6%) of patients were thrombolysed and the most common agent used for thrombolysis was Tenecteplase (52%) followed by Reteplase (17.7%). Approximately 63% of cases had reduced LVEF (<40%) at presentation. All cause in-hospital mortality was 10% and at 30 days, mortality was 12% including 10% in-hospital mortality (Table 1). Almost all the patients were discharged on guideline directed medical therapy: dual antiplatelets (100%), statins (100%), beta blockers (92.5%) and ACE inhibitors/ARBs in 86% (Supplementary Table 1).

**Table 1:** Baseline characteristics of the study group along with in-hospital and 30-day outcomes.

| Variables | Description | In-Hospital |  |  |  | Total | p-value | 30 days outcome |  |  |  | Total | p-value |
| --- | --- | --- | --- | --- | --- | --- | --- | --- | --- | --- | --- | --- | --- |
|  |  | Alive (n=1532) |  | Death(n=168) |  |  |  | Alive (n= 1490) |  | Death(n=210) |  |  |  |
| Age |  | 54.12(±11.79) |  | 57.60(±12.63) |  | 54.46(±11.92) | 0.001 | 54±11.82 |  | 57.72±12.09 |  | 54.46(±11.92) | 0.001 |
| Sex | Female | 278 | 84.2% | 52 | 15.8% | 330 | 0.001 | 271 | 82.1% | 59 | 17.9% | 330 | 0.001 |
|  | Male | 1254 | 91.5% | 116 | 8.5% | 1370 |  | 1219 | 89.0% | 151 | 11.0% | 1370 |  |
| Socioeconomic status | Lower | 198 | 80.8% | 47 | 19.2% | 245 | 0.001 | 194 | 79.2% | 51 | 20.8% | 245 | 0.001 |
|  | Lower Middle | 525 | 92.1% | 45 | 7.9% | 570 |  | 512 | 89.8% | 58 | 10.2% | 570 |  |
|  | Middle | 222 | 89.9% | 25 | 10.1% | 247 |  | 214 | 86.6% | 33 | 13.4% | 247 |  |
|  | Upper | 14 | 100.0% | 0 | 0.0% | 14 |  | 14 | 100.0% | 0 | 0.0% | 14 |  |
|  | Upper Lower | 444 | 91.7% | 40 | 8.3% | 484 |  | 428 | 88.4% | 56 | 11.6% | 484 |  |
|  | Upper Middle | 129 | 92.1% | 11 | 7.9% | 140 |  | 128 | 91.4% | 12 | 8.6% | 140 |  |
| Diabetes Mellitus | No | 1226 | 92.4% | 101 | 7.6% | 1327 | 0.001 | 1197 | 90.2% | 130 | 9.8% | 1327 | 0.001 |
|  | Yes | 306 | 82.0% | 67 | 18.0% | 373 |  | 293 | 78.6% | 80 | 21.4% | 373 |  |
| Hypertension | No | 1067 | 91.5% | 99 | 8.5% | 1166 | 0.006 | 1046 | 89.7% | 120 | 10.3% | 1166 | 0.001 |
|  | Yes | 465 | 87.1% | 69 | 12.9% | 534 |  | 444 | 83.1% | 90 | 16.9% | 534 |  |
| Smoking | No | 939 | 93.9% | 61 | 6.1% | 1000 | 0.001 | 822 | 85.4% | 140 | 14.6% | 962 | 0.002 |
|  | Yes | 593 | 84.7% | 107 | 15.3% | 700 |  | 668 | 90.5% | 70 | 9.5% | 738 |  |
| Physical activity | No | 849 | 88.3% | 113 | 11.7% | 962 | 0.003 | 920 | 92.0% | 80 | 8.0% | 1000 | 0.001 |
|  | Yes | 683 | 92.5% | 55 | 7.5% | 738 |  | 570 | 81.4% | 130 | 18.6% | 700 |  |
| Presentation status | Cardiogenic Shock | 125 | 68.3% | 58 | 31.7% | 183 | 0.001 | 119 | 65.0% | 64 | 35.0% | 183 | 0.001 |
|  | Chest Pain | 1402 | 93.1% | 104 | 6.9% | 1506 |  | 1369 | 90.9% | 137 | 9.1% | 1506 |  |
|  | Heart failure | 5 | 45.5% | 6 | 54.5% | 11 |  | 2 | 18.2% | 9 | 81.8% | 11 |  |
| Heart rate |  | 83.86±18.50 |  | 90.23±21.31 |  | 84.49±18.89 | 0.001 | 83.80±18.50 |  | 89.37±20.83 |  | 84.49±18.89 | 0.001 |
| Systolic BP (mm Hg) |  | 124.18±23.04 |  | 113.15±25.67 |  | 123.15±23.51 | 0.001 | 124.22±23.04 |  | 115.08±25.44 |  | 123.15±23.51 | 0.001 |
| Diastolic BP (mm Hg) |  | 78.00±12.87 |  | 72.40±14.80 |  | 77.48±13.16 | 0.001 | 78.01±12.89 |  | 73.48±14.45 |  | 77.48±13.16 | 0.001 |
| Time since symptoms onset to first ECG |  | 7.64±9.02 |  | 10.39±12.81 |  | 7.90±9.49 | 0.001 | 7.69±9.09 |  | 9.46±11.89 |  | 7.90±9.49 | 0.011 |
|  |  | Alive (n=1532) |  | Death(n=168) |  |  |  | Alive (n= 1490) |  | Death(n=210) |  |  |  |
| (hours) |  |  |  |  |  |  |  |  |  |  |  |  |  |
| Serum creatinine (mg/dl) |  | 1.06±0.44 |  | 1.03±0.35 |  | 1.05±0.43 | 0.439 | 1.05±0.42 |  | 1.07±0.51 |  | 1.05±0.43 | 0.463 |
| Hemoglobin (gm/dl) |  | 13.14±1.73 |  | 12.79±1.74 |  | 13.11±1.73 | 0.013 | 13.13±1.73 |  | 12.97±1.79 |  | 13.11±1.73 | 0.221 |
| Total cholesterol (mg/dl) |  | 165.84±44.67 |  | 162.79±49.52 |  | 165.54±45.17 | 0.407 | 165.85±44.57 |  | 163.31±49.25 |  | 165.54±45.17 | 0.446 |
| LDL (mg/dl) |  | 97.93±35.46 |  | 93.96±39.24 |  | 97.54±35.86 | 0.173 | 97.89±35.42 |  | 95.04±38.82 |  | 97.54±35.86 | 0.281 |
| HDL (mg/dl) |  | 37.11±9.36 |  | 37.25±9.36 |  | 37.12±9.35 | 0.851 | 37.12±9.37 |  | 37.11±9.23 |  | 37.12±9.35 | 0.988 |
| Triglycerides (mg/dl) |  | 158.48±77.12 |  | 156.74±83.26 |  | 158.31±77.72 | 0.782 | 158.71±77.62 |  | 155.48±78.58 |  | 158.31±77.72 | 0.573 |
| Time since symptoms onset to first medical contact | <1 hour | 177 | 94.1% | 11 | 5.9% | 188 | 0.030 | 170 | 90.4% | 18 | 9.6% | 188 | 0.245 |
|  | 1-3 hours | 433 | 92.3% | 36 | 7.7% | 469 |  | 129 | 85.4% | 22 | 14.6% | 151 |  |
|  | 3-12 hours | 735 | 88.7% | 94 | 11.3% | 829 |  | 421 | 89.8% | 48 | 10.2% | 469 |  |
|  | 12-24 hours | 57 | 90.5% | 6 | 9.5% | 63 |  | 54 | 85.7% | 9 | 14.3% | 63 |  |
|  | 1-3 days | 130 | 86.1% | 21 | 13.9% | 151 |  | 716 | 86.4% | 113 | 13.6% | 829 |  |
| Type of MI | Anterior Wall MI | 864 | 90.2% | 94 | 9.8% | 958 | 0.47 | 838 | 87.5% | 120 | 12.5% | 958 | 0.779 |
|  | Inferior Wall MI | 634 | 90.4% | 67 | 9.6% | 701 |  | 618 | 88.2% | 83 | 11.8% | 701 |  |
|  | Lateral Wall MI | 23 | 82.1% | 5 | 17.9% | 28 |  | 23 | 82.1% | 5 | 17.9% | 28 |  |
|  | Posterior Wall MI | 11 | 84.6% | 2 | 15.4% | 13 |  | 11 | 84.6% | 2 | 15.4% | 13 |  |
| Ejection Fraction (%) | < 30 | 91 | 77.1% | 27 | 22.9% | 118 | 0.001 | 89 | 75.4% | 29 | 24.6% | 118 | 0.001 |
|  | 30-40 | 843 | 89.1% | 103 | 10.9% | 946 |  | 807 | 85.3% | 139 | 14.7% | 946 |  |
|  | 40-50 | 564 | 93.7% | 38 | 6.3% | 602 |  | 560 | 93.0% | 42 | 7.0% | 602 |  |
|  | > 50 | 34 | 100.0% | 0 | 0.0% | 34 |  | 34 | 100.0% | 0 | 0.0% | 34 |  |
| Killip Class | I | 1425 | 91.5% | 132 | 8.5% | 1557 | 0.001 | 1387 | 89.1% | 170 | 10.9% | 1557 | 0.001 |
|  | II | 28 | 87.5% | 4 | 12.5% | 32 |  | 25 | 78.1% | 7 | 21.9% | 32 |  |
|  | III | 22 | 68.8% | 10 | 31.3% | 32 |  | 22 | 68.8% | 10 | 31.3% | 32 |  |
|  | IV | 57 | 72.2% | 22 | 27.8% | 79 |  | 56 | 70.9% | 23 | 29.1% | 79 |  |
| Mitral Regurgitation | No | 1389 | 94.6% | 79 | 5.4% | 1468 | 0.001 | 1362 | 92.8% | 106 | 7.2% | 1468 | 0.001 |
|  | Mild | 113 | 66.5% | 57 | 33.5% | 170 |  | 107 | 62.9% | 63 | 37.1% | 170 |  |
|  | Moderate | 22 | 55.0% | 18 | 45.0% | 40 |  | 15 | 37.5% | 25 | 62.5% | 40 |  |
|  | Severe | 8 | 36.4% | 14 | 63.6% | 22 |  | 6 | 27.3% | 16 | 72.7% | 22 |  |

**Abbreviations:** BP: blood pressure; MI: myocardial infarction; ECG: electrocardiograph; LDL: low density lipoprotein; HDL: high density lipoprotein

### In-hospital mortality

A total of six ML models were trained on the training dataset. The models trained were Extra Tree, Random Forest, Multiple Perceptron, Logistic regression, CatBoost, and XGBoost. The performance metrics were sensitivity, specificity, F1-score, accuracy and AUC. Results showed that the Random Forest model had the best performance with a sensitivity of 80%, specificity of 74.0%, F1-score of 78.94%, accuracy of 74.74%, and AUC of 80.83%. For the assessment of model generalization, these models were evaluated on previously unseen datasets. All these models performed well on the unseen dataset with sensitivity and specificity above 65% except CatBoost (Table 2). The F1-scores were also good indicating that the models were not overfitting the data. These models also had high accuracies indicating that the models were making accurate predictions overall. Despite the slight decrease in performance on the unseen dataset compared to the training dataset, these ML models, particularly Random Forest and Multiple Perceptron, exhibited impressive performance across all the metrics, suggesting their potential for generalization to new data.

**Table 2:** Comparison of TIMI score results with Different ML model performance on in-hospital mortality.

| Model | Sensitivity |  | Specificity |  | F1-Score |  | Accuracy |  | AUC |  |
| --- | --- | --- | --- | --- | --- | --- | --- | --- | --- | --- |
|  | Training | Unseen | Training | Unseen | Training | Unseen | Training | Unseen | Training | Unseen |
| TIMI Score |  | 70.0% |  | 64.0% |  | 70.96% |  | 64.70% |  | 70.7% |
| Extra Tree | 81.34% | 65.5% | 82.24% | 83% | 82.16% | 83.79% | 82.11% | 81.43% | 82.14% | 74.35% |
| Random Forest | 83.26% | 80% | 77.36% | 74.0% | 80.26% | 78.94% | 80.35% | 74.74% | 86.96% | 80.83% |
| Multiple Perceptron | 83.42% | 72% | 83.69% | 79% | 83.69% | 81.70% | 83.62% | 78.55% | 81.25% | 76.25% |
| CatBoost | 86.57% | 62.5% | 85.08% | 83.3% | 85.55% | 83.18% | 85.83% | 80.86% | 85.42% | 71.85% |
| XGBoost | 93.87% | 65.5% | 81.72% | 80.67% | 87.48% | 81.36% | 87.57% | 78.22% | 86.80% | 70.38% |
| Logistic Regression | 91.09% | 75% | 74.59% | 70.33% | 82.56% | 76.11% | 82.60% | 71.78% | 81.44% | 71.97% |

**Abbreviations:** AUC: area under the curve; TIMI: Thrombolysis in Myocardial Infarction

Notably, CatBoost excelled in accuracy and specificity but lagged in sensitivity. The Random Forest model with a sensitivity of 80% and specificity of 74% on unseen data demonstrated competence in both positive and negative class predictions. Ultimately, the choice of the best-performing model should be tailored to specific applications considering the relative importance of sensitivity and specificity. The TIMI Score sensitivity was 70%, specificity was 64%, accuracy was 64.7% and AUC was 70.7%. In comparison to the TIMI Score, a traditional clinical risk assessment tool, the ML models, especially Random Forest, outperformed it across all performance metrics. The Random Forest model exhibited higher sensitivity, specificity, F1-score, accuracy, and AUC, indicating superior prediction capabilities for both positive and negative outcomes, as well as overall risk assessment in the context of heart disease (Figure 2). Furthermore, the interpretability of our ML models was determined using ShAP force plots (Figure 3), which provide insights into feature contributions. The features that predicted high risk of in-patient mortality included severe mitral regurgitation, cardiogenic shock, diabetes, low blood pressure and low LVEF. The interpretability analysis using ShAP plots unveiled key predictive factors, contributing to a deeper understanding of the models’ decision-making processes.

**Figure 2:** ROC curve on a training data (A), Unseen data (B) set for six different machine learning models for in-hospital mortality.

**Figure 3:** SHAP based on Random Forest in-hospital mortality. Each point on a feature line is a SHAP value of a sample. The y-axis represents feature ranking in descending order. The x-axis represents SHAP value. The right-side (+ve SHAP value) of the central line indicates the mortality class. A greater +ve SHAP value indicates higher impact on the prediction of the mortality class. Features having red color on the left side (high BMI, ejection fraction more than 35%) are in favor of survival.

### 30-day morality

In order to predict the 30-day mortality, six distinct ML models on a training dataset were used. These models included Extra Tree, Random Forest, Multiple Perceptron, Logistic regression, CatBoost and XGBoost. Among these models, Random Forest emerged as the top performer, achieving remarkable results with a sensitivity of 81.63%, specificity of 78.35%, F1-score of 81.50%, accuracy of 78.82% and an AUC of 78.29%. The remaining models demonstrated lower performance but maintained reasonably satisfactory results, with sensitivities and specificity surpassing the 70% threshold except the XGBoost. The models exhibited strong generalization capabilities when evaluated on an unseen dataset. Most of the algorithms achieved sensitivities of at least 75% and AUCs of at least 78%, indicating their potential for predicting 30-day mortality in patients of STEMI. In comparison to the traditional TIMI score, the ML models, particularly Random Forest, outperformed it in predicting 30-day mortality. The TIMI score exhibited a sensitivity of 63.26% and specificity of 63.91% (Table 3) while ML models, especially Random Forest, showed superior predictive capabilities (Figure 4).

**Figure 4:** ROC curve on a training data (A), Unseen data (B) set for six different machine learning models for 30 days mortality.

**Table 3:** Comparison of TIMI score results with Different ML model performance on 30 days mortality.

| Model | Sensitivity |  | Specificity |  | F1 -Score |  | Accuracy |  | AUC |  |
| --- | --- | --- | --- | --- | --- | --- | --- | --- | --- | --- |
|  | Training | Unseen | Training | Unseen | Training | Unseen | Training | Unseen | Training | Unseen |
| TIMI Score |  | 63.26% |  | 63.91% |  | 63.72% |  | 63.82% |  | 63.59% |
| Extra Tree | 78.27% | 79.59% | 81.49% | 81.79% | 79.49% | 83.56% | 79.89% | 81.47% | 79.98% | 80.79% |
| Random Forest | 84.96% | 81.63% | 78.73% | 78.35% | 82.32% | 81.50% | 81.83% | 78.82% | 79.94% | 78.29% |
| Multiple Perceptron | 78.83% | 75.51% | 91.16% | 86.94% | 83.98% | 86.49% | 85.02% | 85.29% | 83.60% | 80.23% |
| CatBoost | 84.96% | 73.47% | 87.29% | 86.25% | 85.92% | 85.72% | 86.13% | 84.41% | 81.33% | 77.26% |
| XGBoost | 87.47% | 63.27% | 89.50% | 84.88% | 88.33% | 83.25% | 88.49% | 81.76% | 87.58% | 75.87% |
| Logistic Regression | 80.22% | 75.51% | 90.33% | 86.94% | 84.46% | 86.49% | 85.30% | 85.29% | 78.18% | 74.33% |

**Abbreviations:** AUC: area under the curve; TIMI: Thrombolysis in Myocardial Infarction

In order to ascertain the factors predictive of 30-day mortality, ShAP force plots were utilized. The factors predictive of mortality at 30 days included mitral regurgitation on presentation, smoking, cardiogenic shock, diabetes, VSR, Killip class, age, female gender, low blood pressure and a low LVEF. Features such as discharge with aspirin, beta blockers, ACE inhibitors as treatment, and higher LVEF (>40%) were associated with higher survival rates (Figure 5). The ML models, particularly Random Forest, were more effective than traditional TIMI score in predicting 30-day mortality. Use of ShAP force plots identified all the features predictive of mortality at 30 days.

**Figure 5:** SHAP based on Random Forest 30 days mortality. Each point on a feature line is a SHAP value of a sample. The y-axis represents feature ranking in descending order. The x-axis represents SHAP value. The right-side (+ve SHAP value) of the central line indicates the mortality class. A greater +ve SHAP value indicates higher impact on the prediction of the mortality class. Features having red color on the left side (aspirin, beta blocker, ACEi/ARBs, ejection fraction more than 40%) are in favor of survival.

## DISCUSSION

The present study used ML models to identify and predict in-hospital and 30-day mortality in patients presenting with STEMI and compared it with TIMI score. ML model outperformed the TIMI score in identifying patients at high risk for in-hospital and 30-day mortality. CVD remains a major cause of mortality and morbidity in low-and-middle-income countries^16^. In India, 52% of CVD deaths occur before 70 years of age in comparison to 23% in the western populations^17^. Data from the CREATE registry has shown that cardiovascular diseases (CVD) among Indians not only occurs a decade earlier but is more severe^18^. In such resource limited countries, it is important to have accurate risk prediction models to identify high risk populations, triage them and allocate resources wisely.^19^

Traditionally, TIMI risk score which was originally designed and validated among the western population has also been used in the South Asian population to predict 30-day mortality in fibrinolysis eligible patients. These scores have not been well validated in the South Asian substrate which have different ethnicity, varying genetic and risk factor profile. Additionally, TIMI score is limited by using only selected features which identify high risk population group. TIMI score has been known to underestimate mortality risk in those individuals with lower risk which might lead to a delay in initiation of treatment thereby leading to excess avoidable deaths.^20^ Surprisingly, it has also been used among non-thrombolysed patients for which it was never intended to due to non-availability of better risk prediction models. The present study is the first study to develop a ML based score for predicting mortality in STEMI patients and compare it with traditional TIMI score in patients of South Asian ancestry. In the present study, out of all the models trained, Random Forest model performed best for predicting the all cause in-hospital and 30-day mortality. Despite a slight dip in sensitivity in validation set, the model performed very well and was not “overfitted”. The Random Forest model had a high sensitivity of 80% with accuracy of 74% and sensitivity of 81.63% with 78.82% accuracy on unseen data set for predicting in-hospital mortality and 30-day mortality respectively. That makes it apt for practical use in resource limited populous countries like India. In our study, ML based model prediction outperformed TIMI risk score. This finding is in concurrence with that reported in a previous study among the Malaysian population wherein ML model had outperformed TIMI score in STEMI patients^19^. Authors had previously designed a ML based Extra-Tree Classifier model (MERC model) that was able to identify 85% of high-risk patients of STEMI. This significantly impacted the management strategies in the patients of STEMI^21^. However, the previous score required to have the angiographic and PCI details of the patient. These details are often not available because many of the patients do not undergo angiography and PCI. The present score addressed this problem and used only those common features and clinical details that are easily available at presentation and admission for almost all the patients. Using these multiple factors, it calculates the risk of mortality in patients of STEMI. Many such studies from different countries and populations have shown better risk prediction with ML models, but the present study systematically compared this model with traditional TIMI risk score^10,19,22–24^.

Traditionally, TIMI and GRACE risk scores were used extensively for risk stratification in post STEMI patients. TIMI risk score was developed by Morrow and colleagues^3^ from comprehensive multivariable analysis in a population of almost 15000 patients with STEMI from InTIME (Treatment of Infarcting Myocardium Early II) trial. The baseline clinical parameters were estimated as a univariate predictor of mortality of which sixteen parameters were found to be significant predictors of mortality. Of these, ten variables reported a higher predictive value and were included in TIMI risk score where three historical characteristics (diabetes, hypertension, and prior angina) were grouped as a composite variable. The TIMI risk score reported a strong association with mortality at 30 days following STEMI. The prognostic discriminatory capacity of the TIMI risk score was comparable to the full multivariable model. However, traditional risk scores such as TIMI have multiple limitations. TIMI risk score was mainly validated among the Caucasian population. South Asian population consists of diverse ethnicities, varying risk factor profiles and younger age of presentation as compared to the western counterparts as a result the extrapolation of TIMI score to this population group requires validation. Previous studies have attempted to validate TIMI risk score among Asian population with variable results^25^. In one of the larger studies involving more than 40,000 patients, there was only a modest prognostic discriminative value of this score. Infact, the accuracy of TIMI score was worse in approximately 2/3^rd^ of the STEMI patients who did not receive reperfusion therapy. So, the authors suggested cautious use of these scores in various population substrates like the elderly^26^. Exclusion of fibrinolysis ineligible patients, lack of use of LVEF and non-inclusion of other demographic features and presentation profiles are major limitations of this more than two-decade old score. Traditional risk scores such as TIMI that have been developed from statistical methods and Cox proportional regression models have additional limitations as these are based on fixed assumptions centered on the data behavior with limited variables and the need to pre-select predictors at the time of model development. ML algorithms on the other hand are not limited by the number of variables and no prior assumptions are made. These learn from the training data set and predict the individual risk rather than a population-based risk.

The ShAP summary plot in the present study delineated features that significantly impacted mortality and survival. In our study, features such as cardiogenic shock, heart failure on presentation, LVEF < 35%, severe mitral regurgitation, higher Killip class, diabetes mellitus, hypertension, smoking, older age, treatment delay more than one hour, heart rate more than 100 beats per minute, mechanical complications such as ventricular septal rupture and female gender predicted higher in hospital mortality while features such as LVEF >35% predicted favorable outcomes. Features that predicted mortality at 30 days were almost similar with poor LVEF, cardiogenic shock and heart failure on presentation, smoking and diabetes being the important predictors. Whilst on the other hand, LVEF > 40%, use of aspirin, beta blockers, ACEi/ARBs significantly favored survival in this cohort of patient. Lower LVEF is already an established risk factor for higher mortality as reported by sub analysis of NORIN-STEMI registry^27^. Cardiogenic shock, reported in up to 10% of STEMI patients, carries high mortality rates of around 40% at 30 days and 50% at one year ^28^. Landmark trials such as the CULPRIT SHOCK reported that coronary angiography and revascularization of culprit artery can significantly reduce 30 days mortality from 51.6% to 43.3%, further supporting the fact that accurate prediction and triaging of such high-risk patients is of paramount importance^29^.

Diabetic patients have impaired insulin uptake leading to increased triglycerides, lower HDL levels, increased coagulability, impaired fibrinolysis, reduced production of vasodilators like nitric oxide and increased production of vasoconstrictors like endothelin-1 leading to higher incidence of ACS and higher mortality rates^30^. The presence of heart failure on presentation predicts poor prognosis and has been independently linked with three times increased risk of mortality^31^. Nearly a third of the female population present with atypical symptoms and tend to present late. Additionally, females with MI have older age and are less likely to receive evidence-based treatment leading to higher mortality as reported in previous studies^32,33^. Medical therapy comprising beta blockers and ACEi are known to reduce post MI mortality because in such patients there is chronic upregulation of neurohormonal activity in the sympathetic nervous system and renin-angiotensin-aldosterone system (RAAS) that exerts deleterious effect on the myocardium. These drugs are known to downregulate this overactivity and helps in improving left ventricular function and hence, better clinical outcomes^35^. In our model too, STEMI patients who were discharged on these medications showed better clinical outcomes at 30 days of follow-up. ML algorithms have increasingly been used for risk stratification in patients with ACS. ML model comprises of various algorithms with each of them possessing unique advantages tailored to the dataset. Use of multiple ML algorithms shall help in construction of a ML model which would help in better prognostication of patients with STEMI and identify individuals with high risk features who would in all probability have worse outcomes. Additionally, transparency with rationale in ML model decision-making was ensured with use of ShAP and LIME algorithms. The transparency in decision-making enhances the credibility of ML models, aiding healthcare professionals in developing confidence and reliance on the predictions generated by these models.

## Limitations

There are several limitations of this study. This single center study with relatively small number of patients representing only the North Indian population. This data needs to be validated in larger substrates of population with different ethnicities and subgroups. Further, females are underrepresented in this study and results of this study need to be validated in larger female subgroups. Lastly, it is imperative to emphasize that external validation is required to substantiate these findings before definitive statements regarding outcomes can be made.

## Conclusion

Machine learning based models provide good prediction of in hospital and 30 days mortality in a select population. These models are better than the traditional scoring systems like TIMI.

## Data Availability

Data shall be made available following suitable request to the editors

## Abbreviations List

ML: machine learning
STEMI: ST segment elevation myocardial infarction PCI: percutaneous coronary interventions
LIME: local interpretable model-agnostic explanations ShAP: Shapley Additive explanations
NSTEMI: non-ST-segment myocardial infarction
MI: myocardial infarction
LVEF: left ventricular ejection fraction
VSR: ventricular septal rupture
ROC: receiver operating characteristic AUC: area under the curve
CVD: cardiovascular diseases
RAAS: renin-angiotensin-aldosterone system

## Clinical Perspectives

- ML based models had higher sensitivity, specificity and AUC as compared to TIMI risk score to predict in-hospital and 30-day mortality
- Predictors of worse outcomes in post STEMI patients included delayed presentation, presence of risk factors, cardiogenic shock, heart failure and lower LVEF
- In resource limited countries, ML based scores will help in better prognostication, identifying higher risk patients and ensuring appropriate and judicious use of limited resources thereby decreasing hospital stay and cost of health care
- ML based algorithms can be integrated to the electronic medical record systems with which larger subset of patients can be benefitted

---

**Table S1:**
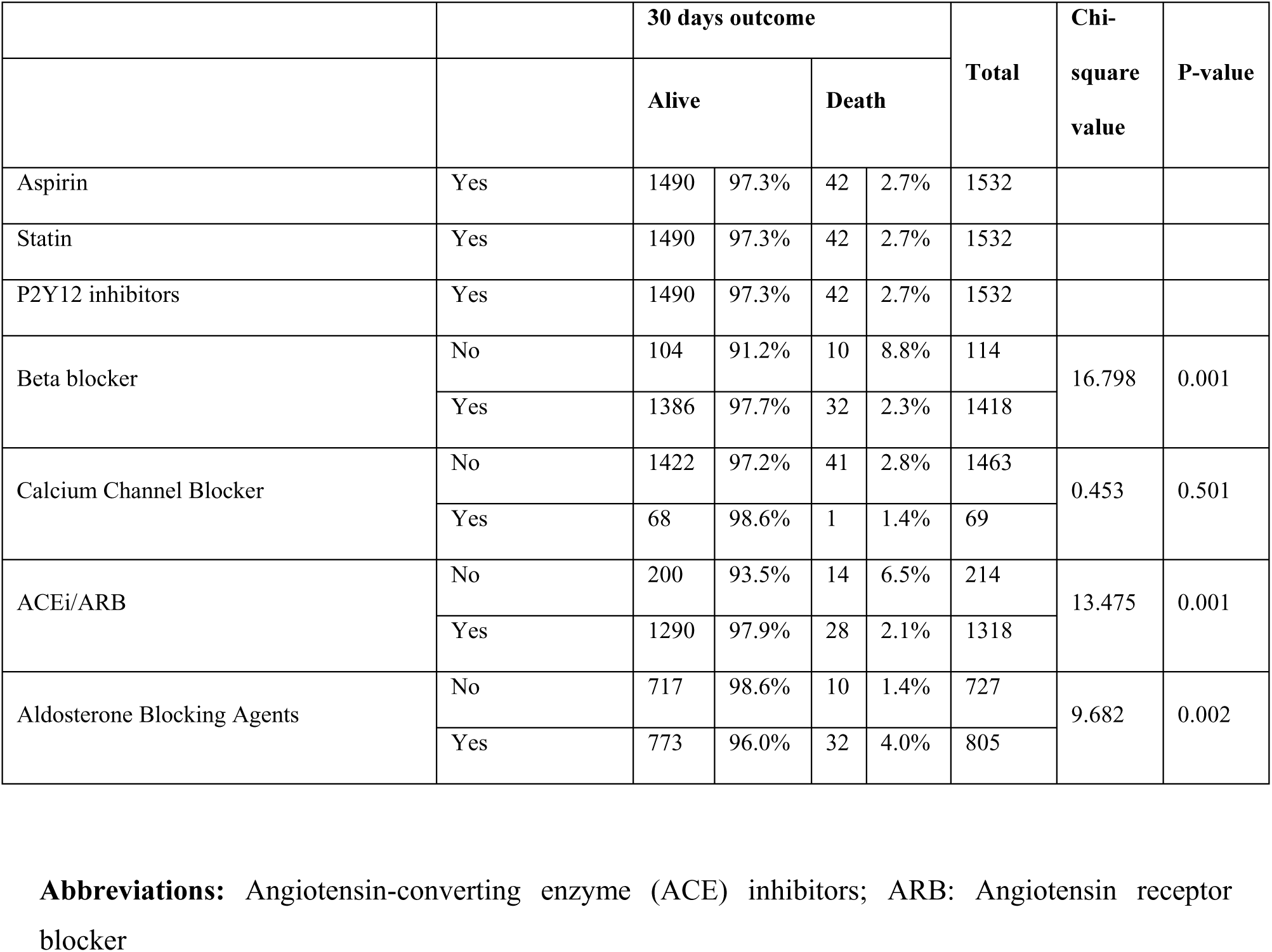
Discharge medications of the study group.

